# Generating Insights Into (Mis)Information About Measles Vaccination in Amsterdam Through Social Listening, the Netherlands, 2024

**DOI:** 10.64898/2026.10.14.26355158

**Authors:** K. M. C. Bisschops, J. C. M. Heijne, C. Lovell, H. L. Coppoolse, M. L. Stein, B. Van den Putte, E. Smit, T. S. Boender

## Abstract

Suboptimal vaccination coverage for Measles, Mumps, and Rubella (MMR) and increases in measles cases in the Netherlands prompted innovation to support informed vaccine decision-making. Through an iterative approach, this pilot implemented and evaluated social listening in the context of a local public health authority, namely Public Health Service Amsterdam. As a pilot study, we generated insights in measles vaccination. First, potential online and offline social listening data sources were identified. Second, in August-September 2024, data on measles vaccination discourse were gathered. Third, quantitative and qualitative (thematic) analyses were applied to generate insights, and risk assessment was conducted to determine their potential impact on health attitudes. We identified twenty sources for social listening. Six were selected for monitored based on feasibility and relevance. These sources were Public Health Service Amsterdam internal observations (public health professionals’ observations, source and contact tracers’ calls, and press inquiries) and three social and news media sources: Coosto, LexisNexis, and Google Trends. Results revealed diverse health beliefs, risk perceptions, concerns, and misinformation shaping vaccination attitudes. Prevalent themes included false claims on effectiveness, causes and effects of measles (vaccination), practical concerns and questions, and positive sentiments. Online and offline social listening enabled insight generation into public health perceptions during an anticipated measles outbreak. This approach is essential for understanding public needs, addressing misinformation, and informing targeted communication during public health challenges.

**Author Summary:** Declining vaccination coverage for measles, mumps, and rubella (MMR) in the Netherlands and concerns about a potential rise in measles cases prompted efforts to better understand public attitudes toward vaccination. To support informed decision-making, we tested a new approach called social listening within the Public Health Service Amsterdam. Social listening combines information from multiple, different sources, both online (e.g. social media, news, and search trends) and offline sources (such as source and contact tracers and reports from public health professionals) to understand what people are saying and exposed to regarding a specific health topic in real time. In this pilot study, we identified and monitored selected data sources during August and September in 2024. We manually analyzed this information to identify common questions, concerns and misinformation about measles vaccination. Combining these insights revealed a wide range of views describing vaccination attitudes, information gaps and themes of distrust and misinformation, but also positive sentiment. This approach provided timely insight into emerging and entrenched public perceptions and showed that social listening can effectively be implemented within a local health service. These findings highlight the potential of integrating social listening into routine public health practice to support responsive and evidence-informed communication strategies.

## Introduction

Social listening, the process of online and offline monitoring, analyzing, and synthesizing conversations has emerged as an approach for systematically monitoring public conversations to identify information needs, emerging concerns, and circulating narratives in near real-time (1). International and national public health organizations, including the WHO, UNICEF, AIRA (Africa Infodemic Response Alliance) US CDC and RKI, subsequently developed guidance, methodology, and examples for applying social listening in public health settings (3–6). Social listening in public health involves systematically collecting and analyzing data from a range of sources, such as social media and other digital or offline channels, to understand public perceptions, concerns, and information gaps. This process focuses on identifying recurring narratives, emerging issues, and patterns of misinformation or distrust. These insights are then synthesized across data sources to generate a broader understanding of the public discourse and inform public health interventions to ultimately enhance the public’s informed decision-making. While frameworks such as the WHO/UNICEF manual (6) provide guidance, social listening remains a young and evolving practice, and approaches are often adapted to context and available resources. The central aim is to translate diverse data into clear, actionable insights that support timely and effective communication and response efforts.

Application of social listening within local public health services remain limited (7). At the Public Health Service Amsterdam (PHSA), a social listening pilot conducted by (8) applied social listening to online sources to capture public discourse on the 2024 HPV vaccination catch-up campaign. The pilot identified several themes that could potentially influence vaccination decision-making and demonstrated that insights could be generated to inform communication strategies. However, the analysis was retrospective and did not include local or interpersonal (“offline”) sources.

Evidence on how social listening can be practically applied within local public health service contexts remains scarce, particularly when integrating larger and more diverse data sets. Local health authorities have direct access to local communities through vaccination programs, schools, daycare centers, and telephone inquiries, enabling them to monitor public concerns in real time. They are responsible for communicating public health guidance, giving them a unique dual role in both observing and influencing public perceptions. This position makes them particularly well-placed to identify emerging concerns and address information gaps promptly, thereby preventing, rather than merely countering, the spread of misinformation (9).

Early 2024, a rise in measles cases and outbreaks elsewhere in the Netherlands (10), coupled with low MMR-vaccination rates in several neighborhoods in region of Amsterdam (11), caused concern among parents and healthcare professionals in Amsterdam. The increased information need (12) made measles vaccination a timely and relevant pilot study to implement social listening at PHSA.

## Aim

The present pilot study aimed to adapt and implement social listening at the local public health service, using measles vaccination in Amsterdam as a pilot study. The question that was addressed using insights was: Which questions and concerns exist around measles vaccination and what misinformation is circulating on this topic in Amsterdam between August and September 2024?

## Materials and methods

### Approach

The social listening framework was implemented in accordance with international guidelines (6, 13) and informed by international examples (3–5, 14) while being adapted to the specific context of the PHSA. We tailored the six steps outlined in the WHO’s guide for social listening (available through https://www.who.int/publications/i/item/9789240075658). The relevance and applicability of each framework step were explored iteratively: adaptation and testing occurred in parallel, with findings from each step informing subsequent adjustments, especially during data collection and analysis. Where possible, the study focused on residents within the PHSA region.

### Data sources inventory

We identified potential data sources and tools relevant for social listening in the context of measles vaccination in the region of Amsterdam, differentiating between three categories of data sources, to include data from: 1) data sources available from PHSA, 2) news media and social media platforms, and 3) data sources from other health institutes. Several factors were tabulated for each potential data source to determine their relevance, expected unique yield, type of data that can be extracted (quantitative or qualitative) and in which format, data extraction frequency, (monetary) costs associated with data extraction, and the extent to which there may be data protection risks or ethical concerns (15). Beyond identifying sources relevant for active monitoring in the present study, the data sources were organized into an inventory to offer an overview of potential sources for future social listening activities (see Table 1 for an overview of identified and included sources).

**Table 1.** Social listening data source inventory, with selection and inclusion for current data analysis (measles vaccination, August - September 2024)

| Category | Identified source | Included in analysis | Reasons for exclusion during analysis |
| --- | --- | --- | --- |
| Public Health Service Amsterdam (PHSA) | Vaccination coverage rates per area in the region of Amsterdam | ✗ | No data on discourse |
|  | Observations from public health professionals | ✗ | No observations on measles during data collection period |
|  | Source & Contact Tracers calls | ✓ | Data included |
|  | Recorded long calls (>6 min) | ✗ | No long calls during data collection period |
|  | Questions asked during live information sessions | ✗ | No information session during data collection period |
|  | Incoming questions/ observations from partners of PHSA | ✗ | No observations on Measles during data collection period |
|  | Responses to targeted campaigns | ✗ | No targeted campaigns |
|  | PHSA Webpage statistics (number of visits, downloads) | ✗ | Effortful, while added value remains unclear |
|  | Notes on phone calls about missed vaccinations | ✗ | No phone calls during data collection period |
|  | 24/7 Press inquiries archive | ✗ | No press inquiries during data collection period |
| Social media and news media | LexisNexis | ✓ | Data included |
|  | Coosto | ✓ | Data included |
|  | Other social listening tools (e.g. Talkwalker, AnswerThePublic) | ✗ | Higher price and lower accessibility for PHSA compared to Coosto and LexisNexis |
|  | Google Trends | ✓ | Data included |
| Other health institutes | Contact point of the National Institute for Public Health and the Environment (RIVM) | ✗ | Access to data could not be arranged in time |
|  | CRIOS data (information from other PHS in the Netherlands (RIVM)) | ✗ | Access to data could not be arranged in time |
|  | *Twijfeltelefoon* (information on vaccination hotline) | ✗ | Access to data could not be arranged in time |

Data sources within PHSA and data sources from other health institutes were inventoried through meetings with stakeholders; i.e. public health professionals working on measles prevention (vaccination), and communication within PHSA and National Institute for Public Health and the Environment (RIVM) and ‘Twijfeltelefoon’. These conversations were held using a semi-structured interview format.

Social media and news media platforms, and social listening tools designed to monitor these, were inventoried through online search and consultations with the PHSA communication department, with the aim of identifying tools already operational within the organization. The Reuters Digital News Report 2024 (16) was used as guidance on which platforms were most used and trusted by Dutch citizens, and to assess whether the selected social listening tools provided coverage of these platforms.

### Data source selection

The data sources selected for active listening were chosen based on unique yield, monetary cost, and availability. No data sources from contacted health institutes could be accessed during the period of data collection as tools to extract and anonymize data were not available on time, and such sources could therefore not be included.

Regarding PHSA sources, observations from public health professionals, the 24/7 press inquiries platform, and source and contact tracers’ phone lines were selected for active listening. To systematically compile relevant information from selected PHSA sources, a standardized reporting form was developed (Appendix S1a and S1b). The form was distributed to observations from public health professionals (e.g. youth nurses, physician assistants) and colleagues involved in source and contact tracing. We requested reports from the PHSA communication department on archived press inquiries related to measles during the data collection period. Several other potential PHSA sources were inactive due to a period of low measles activity in the region of Amsterdam and the summer holiday break. For instance, no measles-related information sessions were held, and no targeted campaigns were conducted during the summer holidays.

The selection of Coosto, LexisNexis, and Google Trends as social and news media tools was guided by considerations of cost and accessibility as well as the aim to minimize unnecessary additional work and to leverage existing systems. To ensure consistency across results from social and news platforms, the search query “Measles (vaccin* OR infection OR outbreak)” (Originally in Dutch: “Mazelen (vaccin* OR infectie OR uitbraak)”) was applied. LexisNexis provided near-daily email updates on relevant news items, which were manually reviewed and coded for publication date, source, platform type, and relevance to the research question. Coosto was used to collect both qualitative and quantitative data from a broad range of sources, including social media platforms (e.g. X, Facebook, Instagram), blogs, forums, and news websites, providing insight into both the public discourse and engagement. Due to time constraints regarding the manual coding of all items, the top 100 most viewed posts and articles per month were included. Google Trends data were used to assess search inquiry frequency related to ‘Measles’. The online sources covered the entire Netherlands, as data specific to the Amsterdam region was limited and the location of online content was not always identifiable.

### Data collection

Data was collected between August 1 and September 30, 2024. This timeframe was selected based on early signals in July indicating potential measles cases in the Amsterdam region, prompting monitoring in anticipation of a possible outbreak, as well as the timing of the pilot. Notably, no measles outbreaks or clusters occurred during this period in Amsterdam. Although no outbreaks occurred and some sources yielded limited data, this period allowed monitoring baseline public discourse and early signals that could inform future communication.

### Integrated data analysis

Separate analysis plans were developed for each source to account for differences in data format and content, though all sources providing qualitative data were ultimately analyzed using manual thematic analysis. Two researchers (KMCB and TSB) conducted analyses independently for each qualitative data source: Coosto, LexisNexis, and source and contact tracers telephone calls.

Initially, all data were summarized to ensure familiarity with content and context, followed by two rounds of independent coding in a spreadsheet. Data, including social media posts, online reactions, news articles, information pages, and telephone call notes, were coded for recurring themes and uniquely relevant issues. An integrated analysis combined themes across sources through manual coding and content description. Data was conceptually categorized into three types: relevant discourse, which reflects public perceptions and is the focus of this pilot; contextual insights, providing background or explanatory information but not representing public discourse directly; and noise, which includes irrelevant or off-topic data. Only the relevant discourse was carried forward into the thematic analysis and risk assessment, while contextual insights and noise were excluded.

Finally, to validate the themes for face validity, the results were presented to a group of vaccination and health-related communication experts affiliated with PHSA, who reviewed and confirmed the relevance, accuracy and exhaustiveness of the identified themes.

### Risk assessment

Themes identified were classified using a risk assessment matrix, as shown in Table 2, based and building on CDC’s State of Vaccine Confidence Insight Reporting System and the WHO manual(6) .

**Table 2.**
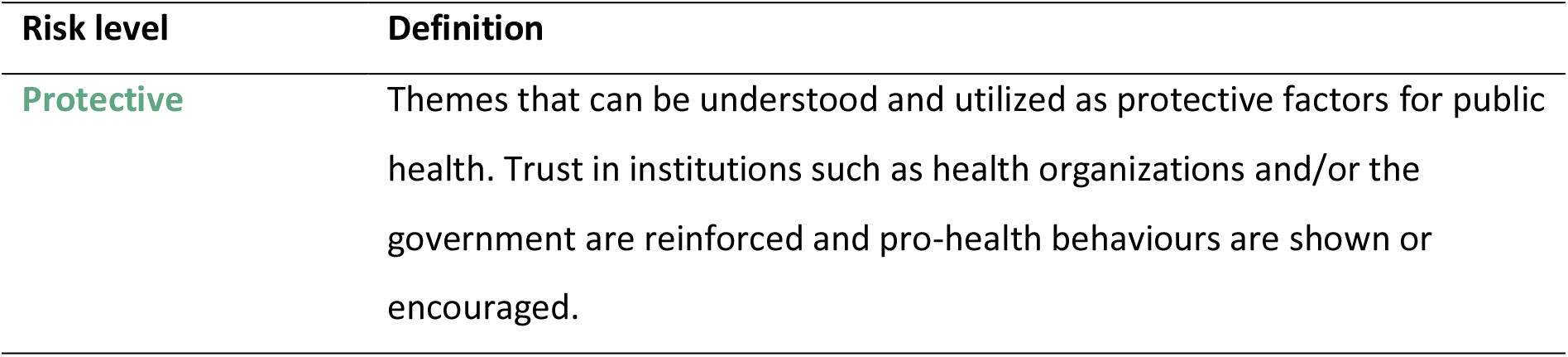
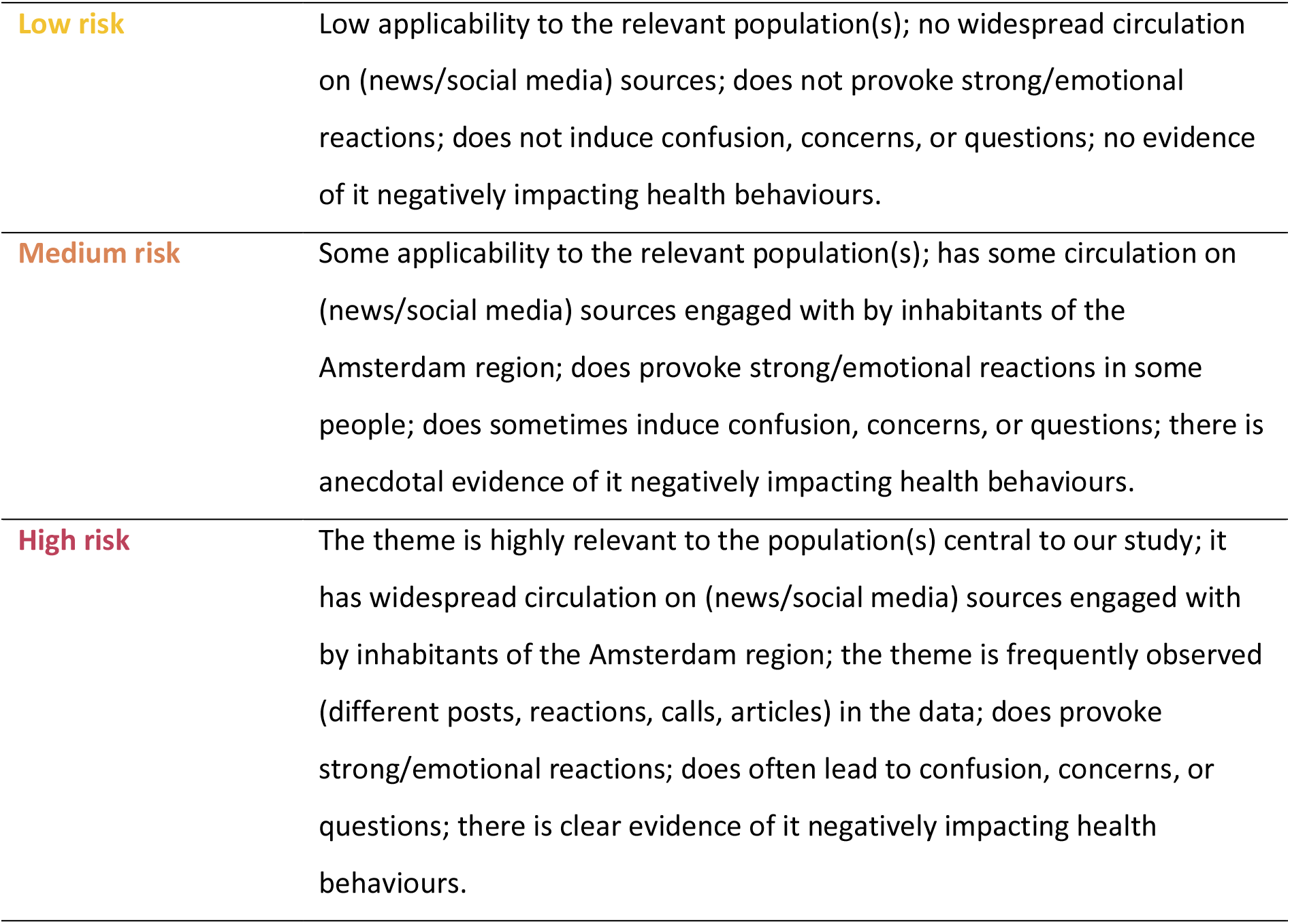
The public health risk assessment matrix used to classify themes identified by social listening

| Risk level | Definition |
| --- | --- |
| Protective | Themes that can be understood and utilized as protective factors for public health. Trust in institutions such as health organizations and/or the government are reinforced and pro-health behaviours are shown or encouraged. |
| **Low risk** | Low applicability to the relevant population(s); no widespread circulation on (news/social media) sources; does not provoke strong/emotional reactions; does not induce confusion, concerns, or questions; no evidence of it negatively impacting health behaviours. |
| **Medium risk** | Some applicability to the relevant population(s); has some circulation on (news/social media) sources engaged with by inhabitants of the Amsterdam region; does provoke strong/emotional reactions in some people; does sometimes induce confusion, concerns, or questions; there is anecdotal evidence of it negatively impacting health behaviours. |
| **High risk** | The theme is highly relevant to the population(s) central to our study; it has widespread circulation on (news/social media) sources engaged with by inhabitants of the Amsterdam region; the theme is frequently observed (different posts, reactions, calls, articles) in the data; does provoke strong/emotional reactions; does often lead to confusion, concerns, or questions; there is clear evidence of it negatively impacting health behaviours. |

Two researchers (KMCB and TSB) independently assessed the risk of each identified theme by measuring its spread (e.g., number of online views or phone calls), interpreting how it might affect health behaviours and attitudes toward health institutions, and discussed the assessment with the project team.

### Data protection and ethics

According to applicable regulations, formal ethical review was not required for this study, as it relied exclusively on publicly available data and routinely collected, anonymized operational data, and did not involve interventions on human subjects. All tools developed for anonymous data collection were designed to exclude any personal identifiable information. Specifically, source and contact tracers phone calls were continuously archived as part of standard practice, and observations from public health professionals were documented during regular meetings. A standardized form was distributed to the nurses and source and contact tracers’ employees to collect anonymous data specifically relevant to this study. For online sources, any identifiers potentially linking data to specific individuals (e.g., account names) were removed during the data cleaning process to ensure anonymity. In cases where data sources contained other privacy-sensitive information or posed data sharing considerations, data were collected in aggregated form. The raw data were securely stored within the protected ICT infrastructure of PHSA. We followed WHO’s ethical guidance for social listening (15) to ensure that our activities were in concordance with the protection of and respect for human rights.

## Results

### Descriptive statistics

PHSA source and contact tracer employees recorded information on two phone calls in August, from which themes with a medium risk were identified. Public health professionals did not observe or receive any questions regarding measles vaccination, nor did the communication department receive any press inquiries during the data collection period.

Coosto provided a total of 200 publications (100 per month of monitoring) for analysis; including posts, reactions, articles, and other content drawn from social media platforms, news outlets, forums, and blogs or informational websites. Coosto data yielded the most diverse set of themes, encompassing all risk categories described earlier.

LexisNexis yielded 247 results consisting of printed or digitally published news articles, all of which were included in the analysis. Some articles were published across different platforms; after accounting for duplicates, 143 unique titles were identified in LexisNexis for August and September, originating from 107 distinct news platforms.

### Identified themes

Of the 16 themes identified (Table 3), five were classified as high risk, three as medium risk, four as low risk, and four as protective. Interestingly, 15 out of 16 themes describing the public discourse originated from X.

**Table 3.** Overview of identified themes with associated risk levels, data sources, and engagement frequencies using social listening for measles-vaccination, August-September 2024, Amsterdam region, the Netherlands.

| # | Identified theme | Risk level | Sources | Engagement |
| --- | --- | --- | --- | --- |
| 1 | Doubts about the effectiveness and safety of the measles vaccine | High | X | 53,893 views |
| 2 | “Vaccine propaganda” and the autism narrative | High | X | 21,098 views |
| 3 | A preference for natural immunity | High | X | 19,255 views |
| 4 | COVID-19 is said to cause immunodeficiency and thereby indirectly cause measles | High | X | 14,123 views |
| 5 | Misrepresentation of historical data (graph) | High | X | 6,090 views |
| 6 | Questions about measles-risk in the context of suspected measles cases | Medium | Source and<br>Contact tracers<br>phonelines | 2 calls |
| 7 | Vaccines as a suspected source of disease | Medium | X | 101,444 views |
| 8 | Minority groups blamed for outbreaks | Medium | X | 7,895 views |
| 9 | Concerns about the consequences of a measles infection | Low | X | 52,731 views |
| 10 | Uncertainty about the duration of protection from vaccination | Low | X | 15,326 views |
| 11 | Concerns about combining vaccines | Low | X | 1,666 views |
| 12 | Distrust in reporting on infectious disease outbreaks | Low | X | 21,381 views |
| 13 | Sharing positive experiences and gratitude for vaccination | Protective | X | 209,789 views |
| 14 | Expression of importance of vaccination | Protective | X | 8,307 views |
| 15 | Sharing accurate health information regarding vaccination | Protective | X | 1,377 views |
| 16 | Criticism of people who do not vaccinate (their children) | Protective | X | 10,829 views |

High-risk themes represented attitudes and behaviour to avoid vaccination and to increase distrust in health authorities and vaccination practices or an apparent intention to influence these in others. These themes were often widespread (garnered many views or appeared frequently in the dataset) and occurred in the data a multitude of times (i.e., in multiple separate discussions and posts).

Medium-risk themes reflected content that combined expressions of uncertainty, misattribution of measles causation, and blaming of minority groups, representing potentially harmful discourse but with less intensity or reach than the high-risk themes.

Low-risk themes primarily reflected concerns and questions about measles vaccination, rather than strongly negative sentiments or inaccurate information. For example, a quote (translated from Dutch) demonstrating concern within this category is: ‘[…] *Do the risks of vaccination concern only the measles vaccine, while multiple and combined vaccines are administered during a single vaccination round, for which no safety studies are available?*’

Analysis revealed four protective themes. Even though all protective themes represent positive sentiment regarding measles vaccination, the fourth (Theme 16, criticism of vaccine refusers) is more negative in tone than the first three (Themes 13, 14, and 15), as it attempts to stigmatize people rather than share accurate and positive information. The following (translated) quote demonstrates this negative labeling of vaccine refusal: “*He was already known as an anti-vaxxer, even going so far as to claim that autism was caused by [vaccinations] (including those against [measles]). Not far-right, but extremely crazy.*”

These insights, along with contextual insights, noise and trends, were combined and summarized in detail in an infodemic insights report (Appendix 2). This report is disseminated among health professionals within PHSA.

## Discussion

### Summary

This study piloted social listening within a local public health service to generate insight into the measles vaccination discourse during a period of anticipated measles cases in the region of Amsterdam, the Netherlands. Although no measles cases were reported during the study period, social listening activities and integrated analysis generated actionable insights originating primarily from social media. High risk themes reflected misinformation and narratives of distrust that included doubts about the safety and the effectiveness of the measles vaccine, the autism narrative (17), and an information gap on the harmfulness of measles infection. Medium risk themes included questions about measles-risk in case of suspected measles cases, blaming of minority groups for outbreaks, and concerns about the consequences of vaccines. Low risk themes included distrust in official reports on measles cases, concerns about combining vaccines, and uncertainty about the duration of vaccine protection. Protective themes included defending vaccines, sharing of accurate health information, and expressing gratitude for vaccination efforts.

### Strengths and limitations

This study is the first pilot of social listening in the context of a local public health service in the Netherlands. It combined external social listening tools with internal local health information streams. The iterative approach and independent manual thematic coding, along with structured and data informed risk assessment specifically adapted to the data sources available and the context of PHSA, enhanced the robustness and validity of the findings. Despite these strengths, three methodological considerations should be noted.

First, data source availability was limited by the absence of a measles outbreak in the Amsterdam region, with most identified themes originating from X, which may limit representativeness. Subsequently, the discourse identified was heavily dominated by narratives on social media platform X, which may have resulted in a skewed representation of public opinion. Louder and more polarized voices, such as anti-vaccine or activist communities, were likely overrepresented (18).

Second, we were unable to include private social media groups and video platforms. Future studies should incorporate these sources, as they likely host relevant vaccination-related discourse and are increasingly used by young adults to access health information (19). It is worth noting that access to social media data has become increasingly restricted, even though EU legislation such as the Digital Services Act provides a framework to improve transparency and enable qualified researchers to request platform data (20), which may shape the feasibility of future online monitoring of health-related public discourse.

Third, although online monitoring provides valuable insights, it inherently misses individuals with little to no online activity. Some populations rely primarily on interpersonal sources for health information, such as community centers, personal networks, or contact with health services (of which only the latter was included in this study) (21, 22). By incorporating local public health service data, we also captured some, but very limited offline signals. Future research could further enhance representativeness by incorporating additional offline key figures and community networks, providing a more complete understanding of public concerns and misinformation.

### Comparison with existing literature

Although this study was characterized by a unique context (i.e., during a cold phase, within a local public health service in the Netherlands), the findings were largely consistent with international experiences. For example, a small pilot study on social listening about measles during an outbreak in Moldova (23) found, similar to our study, that identified themes reflected a lack of knowledge about the risks of measles, distrust in authorities, and a variety of political and ideological views. The similarity in findings may partly reflect the fact that, due to the absence of a local outbreak, our analysis was largely limited to social media data. This aligns social listening more closely with earlier social media analyses in this context, although social listening is generally more signal-driven compared to the more measurement-driven nature of traditional social media analysis (e.g. 24). A key strength of social listening at PHSA is that it also allows monitoring of local offline sources, which can provide additional insights during periods of higher disease activity.

## Interpretation

This study demonstrates that social listening can be implemented at the local level. Two key lessons learned emerge from the findings. First, the findings showed that social listening during a period of low disease activity captures a meaningful segment of public discourse, characterized by persistent misinformation, information gaps and varying levels of concern. This demonstrates the proactive value of social listening: it can detect emerging information gaps in a cold phase and identify persistent misinformation before they escalate.

Second, more nuanced perspectives were less apparent, possibly because measles outbreaks in other areas of the Netherlands did not spread to the Amsterdam region. Consequently, local offline sources, which might provide more additional insights during periods of higher disease activity, remained largely inactive. This suggests that online social listening during periods of low disease activity primarily captures expressed and more intense discourse, rather than the full spectrum of beliefs and concerns present within the broader population. It is important to note that the current findings are specific to a period of low disease activity, and the general sentiment could potentially shift during outbreaks. Accordingly, social listening should be interpreted in the broader epidemiological context, and complementary data sources may be needed to capture subtler or less vocal public concerns.

### Implications

The practical implications of this study are multifold. Social listening can help local public health authorities in the early detection of emerging themes, identifying persistent misinformation and genuine public questions and concerns. These insights can be translated into actionable communication strategies, such as improving the accessibility of accurate information on official websites and social media channels, shaping FAQ materials, informing public information sessions, and tailoring messaging to communities most affected by misconceptions to support informed decision-making.

However, some identified themes reflect deeper distrust in authorities and entrenched misinformation, which require more comprehensive approaches (see for exa 25) beyond improving information access or communication alone. In these cases, strategies such as prebunking, aimed at preventing the spread of false narratives, and debunking, targeting existing misinformation, and interventions or actions aimed at improving trust (26, 27) are more appropriate. Our follow-up study will examine the effectiveness of prebunking and debunking, with a particular focus on how their timing can be optimized to maximize impact. Collaborating with trusted community leaders may also strengthen trust while enhancing both monitoring efforts and access to accurate information.

The range of identified risk levels, from high to low, as well as the presence of protective themes, underscores the multidimensionality of public attitudes toward measles vaccination. These findings emphasize the need for targeted and context-sensitive communication strategies. Social listening can play a key role in this process by enabling public health authorities to better understand and respond to this complex landscape (1).

## Conclusions

The current study in the Netherlands demonstrated that social listening can be effectively adapted and implemented to the context of a local public health service. The approach provided insight into diverse, ongoing conversations and information dynamics at times when broader public discussion on measles vaccination was largely absent. It confirmed that social listening may detect information gaps early on and identify narratives during a cold phase that may warrant attention. In addition, it has the potential to prevent misinformation spread through providing accurate information, improving communication strategies, and debunking and prebunking when misinformation is detected in the discourse.

## Supporting information

Ethical statement, not in manuscript

## Data Availability

The minimal dataset underlying the findings will be deposited in a publicly accessible repository (e.g., Zenodo / OSF / Figshare) upon acceptance of the manuscript. The data will be available under an open license.

## Author contributions

TSB conceived and coordinated the pilot and KMCB and TSB established social listening at the Public Health Service Amsterdam. KMCB held stakeholder interviews. KMCB, CL, HC and MS inventoried data sources. KMCB performed data cleaning and structuring. KMCB, CL and HC oversaw data collection. KMCB and TSB conducted thematic analysis. KMCB wrote infodemic insights report. KMCB wrote the first draft of the manuscript, together with TSB, JCMH, BP and ESS, who provided input on the development and interpretation of the work. All authors critically revised the manuscript and approved its final version.

## Acknowledgements

The authors thank Hedwig te Molder, Matthijs Lambooij, Udi Davidovich, Marco Hol, Maarten Schim van der Loeff, Lieke van Houtum and Linda Kreemers for their valuable contributions to the conceptualization and methodology of the study, for providing guidance during its conduct, and for assisting in the evaluation and interpretation of the results. The authors also acknowledge their support in the dissemination of the findings. The authors further thank Bastiaan Buesink for valuable advice on data collection tools and for providing and curating part of the dataset used in this study.

## Funding

Funded by the Dutch Organization for knowledge and innovation in health, healthcare and well-being (ZonMw): 10710032310026.

## Appendices

**S1a appendix.** Standardized form distributed among employees collecting data for the pilot

| Date | Question/observation | Age group | Type | < Other, namely: | Area (optional) |
| --- | --- | --- | --- | --- | --- |
| dd-mm-yyyy | Short summary | Select option | Select option |  | Select option (optional) |

**S1b appendix.** Description of the columns from the standardized (1a) form, also distributed to employees collecting data

| Column | Explanation |
| --- | --- |
| Date | Date of the (telephone) conversation/observation dd-mm-yyyy. |
| Question/observation | Short summary of the question/observation. |
| Age group | Select one of the age groups (<14 months, 0-4 years, 5-18 years, adult, adult born between 1965 and 1975). This concerns the age group of the person the question is about. |
| Type | Select type of caller (citizen, health care professional, school, daycare, other). In case of 'Other', specify in the next column. |
| < Other, namely: | When 'Other' was selected, specify the type of caller (function), otherwise leave empty. |
| Area | Optional: select the area in or near Amsterdam relevant to the question/observation. |

## S2 appendix. Infodemic insights report

Please view the anonymized supplementary insights report

## Notes

### Competing Interest Statement

The authors have declared no competing interest.

### Author Declarations

The Compliance and Privacy Officer of Public Health Service (PHS, or GGD in Dutch) waived ethical approval for this work. Their statement: 30 July 2026 To: MedRxiv editorial team Subject: Ethics declaration To whom it may concern, The study 'Generating Insights Into (Mis)Information About Measles Vaccination in Amsterdam Through Social Listening, the Netherlands, 2024' (MedRxiv ID: MEDRXIC/2026/355158) was conducted as part of routine public health practice within the statutory responsibilities of the Public Health Service Amsterdam under the Dutch Public Health Act (Wet publieke gezondheid, Bulletin of Acts and Decrees 2008, No. 460). The study used publicly available online information and aggregated information collected through routine source and contact tracing activities. During the analysis of publicly available online content, author/usernames and URLs were temporarily available to the researchers solely for the purpose of understanding the context of individual messages. These data were not retained in the research dataset and were not included in the manuscript or reported results. The information obtained through source and contact tracing did not contain names, contact details or other information that could be used to identify individual persons. The information consisted of general questions or concerns, broad age categories and caller type. No information enabling identification of individual persons was included in the analyses or reporting. Appropriate measures were taken to minimise the use of identifiable information and to protect the privacy and confidentiality of the data. The research results were reported in aggregated form and do not contain information that could identify to individual persons. The study did not involve medical interventions. Consequently, formal ethical review under the Dutch Medical Research Involving Human Subjects Act (Wet medisch-wetenschappelijk onderzoek met mensen; WMO) was not required. A formal WMO review was therefore deemed unnecessary in accordance with the Dutch Medical Research Involving Human Subjects Act. The Public Health Service Amsterdam confirms that appropriate measures were taken to ensure privacy protection and the confidentiality of the data used in this study. The privacy aspects of the study have been retrospectively assessed by the Compliance and Privacy Officer of the Public Health Service Amsterdam. Based on this assessment, the compliance and privacy officer confirm that appropriate privacy and confidentiality safeguards were applied and that no additional institutional privacy review was required for this study. Sincerely, Compliance and Privacy Officer GGD Amsterdam

